# The power and limitations of genomics to track COVID-19 outbreaks: a case study from New Zealand

**DOI:** 10.1101/2020.10.28.20221853

**Authors:** Jemma L Geoghegan, Jordan Douglas, Xiaoyun Ren, Matthew Storey, James Hadfield, Olin K Silander, Nikki E Freed, Lauren Jelley, Sarah Jefferies, Jillian Sherwood, Shevaun Paine, Sue Huang, Andrew Sporle, Michael G Baker, David R Murdoch, Alexei J Drummond, David Welch, Colin R Simpson, Nigel French, Edward C Holmes, Joep de Ligt

## Abstract

**Background:** Real-time genomic sequencing has played a major role in tracking the global spread and local transmission of SARS-CoV-2, contributing greatly to disease mitigation strategies. After effectively eliminating the virus, New Zealand experienced a second outbreak of SARS-CoV-2 in August 2020. During this August outbreak, New Zealand utilised genomic sequencing in a primary role to support its track and trace efforts for the first time, leading to a second successful elimination of the virus.

**Methods:** We generated the genomes of 80% of the laboratory-confirmed samples of SARS-CoV-2 from New Zealand’s August 2020 outbreak and compared these genomes to the available global genomic data.

**Findings:** Genomic sequencing was able to rapidly identify that the new COVID-19 cases in New Zealand belonged to a single cluster and hence resulted from a single introduction. However, successful identification of the origin of this outbreak was impeded by substantial biases and gaps in global sequencing data.

**Interpretation:** Access to a broader and more heterogenous sample of global genomic data would strengthen efforts to locate the source of any new outbreaks.

**Funding:** This work was funded by the Ministry of Health of New Zealand, New Zealand Ministry of Business, Innovation and Employment COVID-19 Innovation Acceleration Fund (CIAF-0470), ESR Strategic Innovation Fund and the New Zealand Health Research Council (20/1018 and 20/1041).

## Main Text

Only twelve days after the novel coronavirus SARS-CoV-2 was identified, a genome of the virus was first published^1^. This information was pivotal to the subsequent, rapid, development of diagnostic tests and identification of potential treatments^2,3^. Since then, as of 25 September 2020, over 110,000 genomes of SARS-CoV-2 have been shared publicly^4^. The underlying genome sequencing has occurred so quickly that, for the first time during an infectious disease outbreak, it has enabled virological and epidemiological data to be integrated in real time^5^. Analysis of these data has played an important role in informing the COVID-19 response by tracking the global spread and evolution of SARS-CoV-2, including identification of the number, source, and timing of introductions into individual countries. This has led to a greater understanding of COVID-19 outbreaks around the world^6-13^.

Of the 185 countries that have reported positive cases of COVID-19 to the World Health Organization^14^, 60% (n=112) have sequenced and shared SARS-CoV-2 genomes on the GISAID database^4^ (September 2020). This immense global sequencing effort has facilitated ongoing genomic surveillance of the pandemic, including monitoring viral genetic changes of interest^12^, which has informed public health responses^15-18^. Nevertheless, the number and proportion of positive cases sequenced, and genomes published, varies dramatically between countries and over time (Figure 1). The COVID-19 Genomics UK (COG-UK) consortium, for example, has led to the UK being the most represented sampling location, totalling over 42,000 genomes and comprising 39% of the global data set despite recording only 1% of the world’s positive cases (n = 412,241). Conversely, SARS-CoV-2 genomes sequenced in India represent just 3% of the global data set but 18% of the world’s total reported cases (n = 5,646,010).

**Figure 1.**
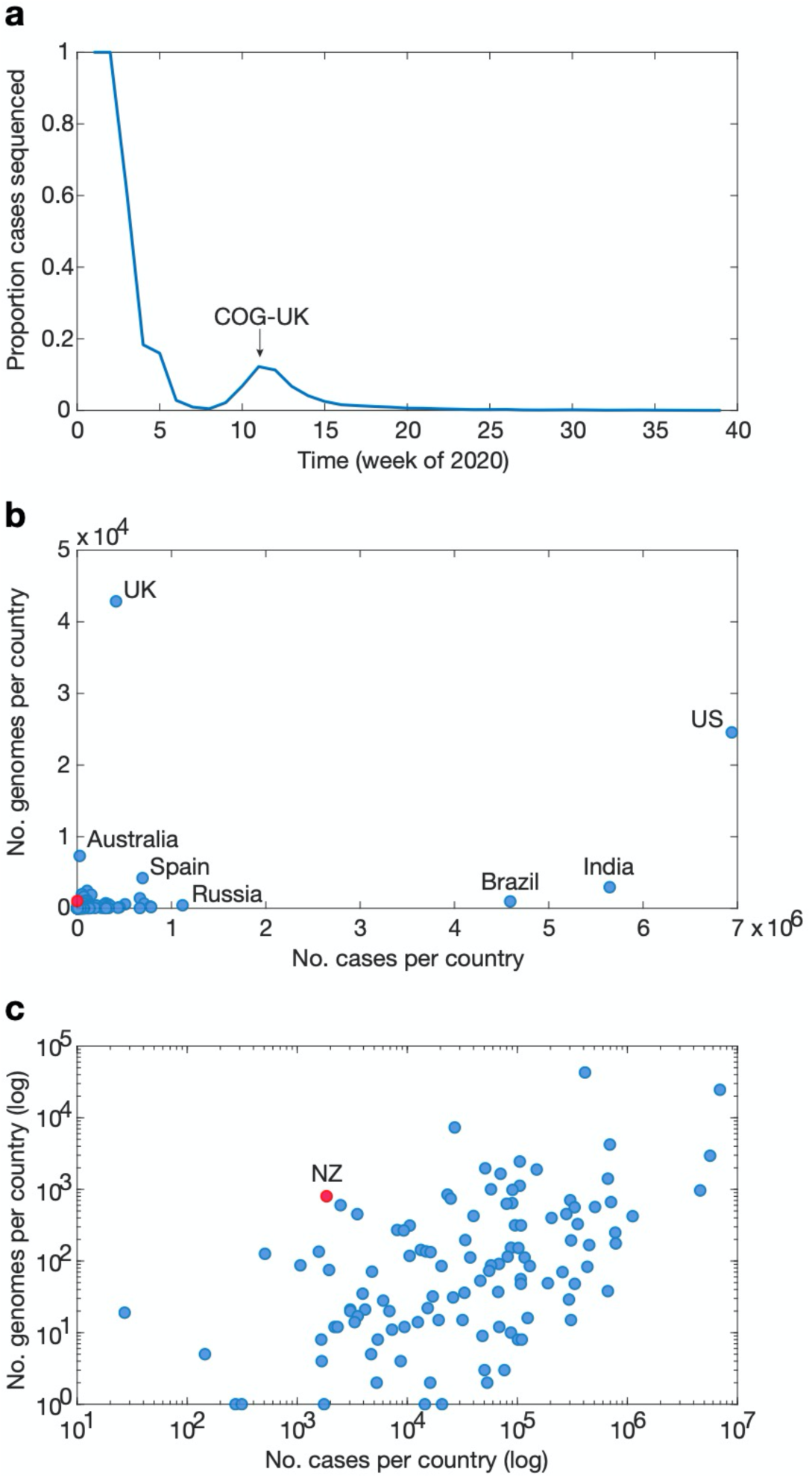
**a** Proportion of global cases sequenced and shared on GISAID between December 2019 to September 2020, where the second mode was largely driven by the COG-UK consortium as illustrated; **b** Number of genomes sequenced and number of reported cases per country (New Zealand is represented in red) on a linear scale; **c** Number of genomes sequenced and number of reported cases per country on a logarithmic scale.

We show here that such disparities in sequencing effort can have major implications for data interpretation and must be met with careful consideration. Real-time sequencing of SARS-CoV-2 genomes has, however, had particular utility in tracking the re-emergence of the virus in New Zealand. By June 2020 New Zealand had effectively eliminated COVID-19 in the community and positive cases were limited to those linked to managed quarantine facilities at the border^7,13,19^. Following over 100 days with no detected community transmission of COVID-19, four new cases emerged on August 12, 2020 with no apparent epidemiological link to any known case. We compared the genomes of these cases to sequenced cases from both New Zealand’s first wave and those in quarantine facilities and, again, found no link. The vast majority of available sequence data from cases in New Zealand’s quarantine facilities were of different virus lineages than that of the August 2020 outbreak. However, this observation was of limited value given that only 42% of cases in those quarantine facilities had adequate viral RNA for successful genomic sequencing. In order to determine the likely origins of this outbreak we compared genomes from the new community outbreak to the global data set.

An initial genomic sequence analysis found that the re-emergence of COVID-19 in New Zealand was due to a SARS-CoV-2 from lineage B.1.1.1^20^. Of the countries that have contributed SARS-CoV-2 data, 40% had genomes of this lineage. Remarkably, 85% of B.1.1.1. genomes were from the UK and were generated between March – September 2020, the vast majority of which were sampled during the first wave of disease in the UK (Figure 2). Phylogenetic analysis of the most recently sampled B.1.1.1. genomes (1,996 of 4,544) identified genomes sampled from Switzerland, South Africa and England in August as the most likely to be contained within the sister clade (Figure 2): these were the closest sampled genomic relatives of the viruses associated with New Zealand’s August 2020 outbreak (Supplementary Figure 1). Additional Bayesian analysis estimated that the outbreak originated 11 days before the first transmission event, with a 95% highest posterior density (HPD) of 0 – 28 days. We also estimated that the first transmission event in the outbreak occurred between 26 July – 13 August 2020 (95% HPD; mean date of 5 August). Epidemiological data showed that two confirmed cases linked to the outbreak had a symptom onset date of 31 July, although the most probable sampled genomes within the sister clade were sampled later, between 6 August – 28 August. Hence, is unlikely that the currently available global genomic data set contains the source of this outbreak.

**Figure 2.**
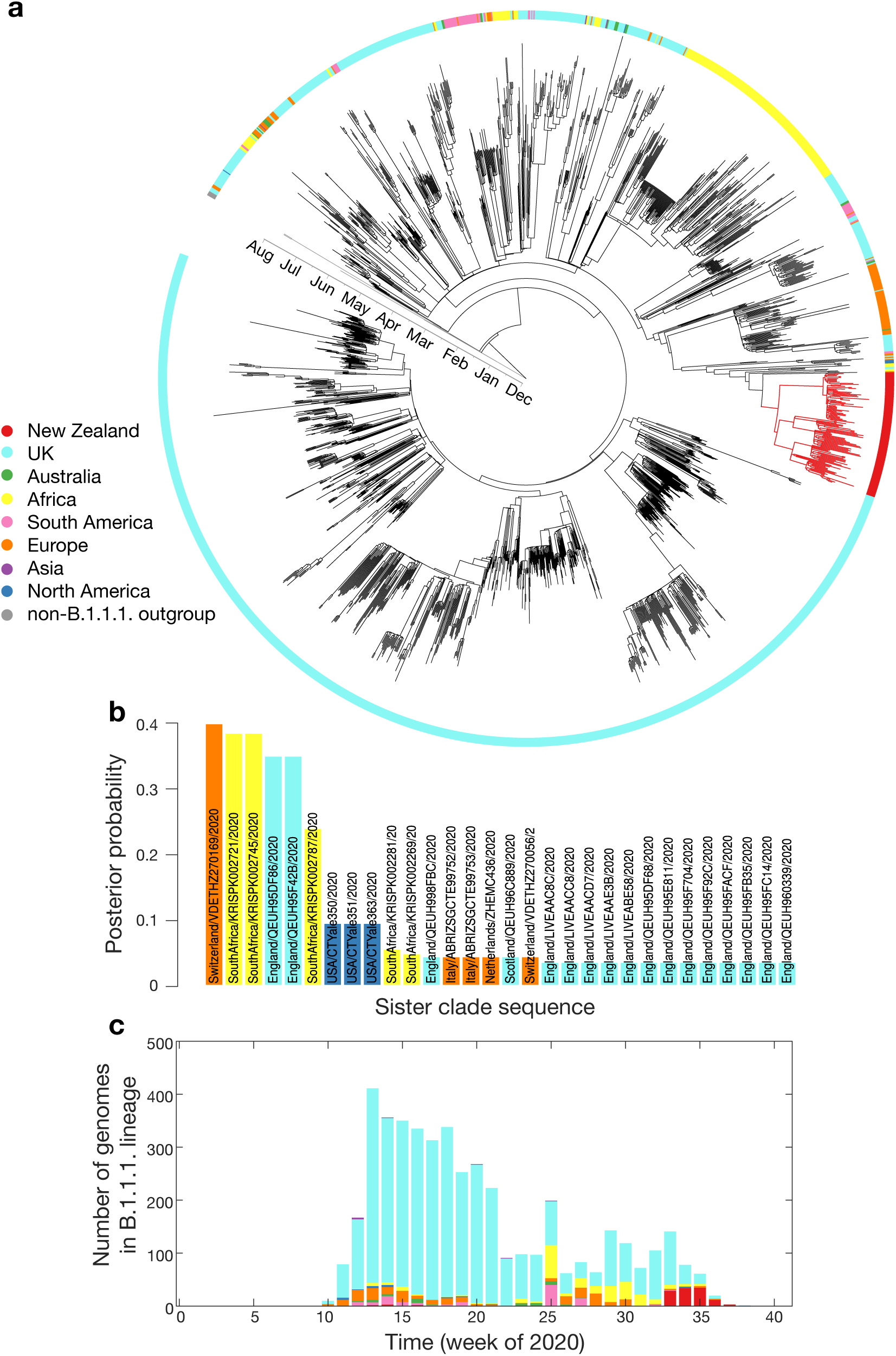
**a** Maximum clade credibility phylogenetic tree of 2,000 subsampled global genomes (1,996 most recently sampled B.1.1.1. plus four non-B.1.1.1. used as an outgroup) with an outer ring coloured by sampling region; **b** Posterior probability of genomes within the sister clade to New Zealand’s August outbreak, colour-coded by sampling location; **c** Proportion of genomes within lineage B.1.1.1. in the global data set over time, colour-coded by sampling location.

Genomic epidemiological analysis on the possible origins of New Zealand’s re-emergence was found to be inconclusive, likely due to missing genomic data within the quarantine border facilities as well as in the global data set. A glimpse into the genomic diversity likely omitted from the global data set can be seen in the genomes sequenced in New Zealand from positive quarantine cases which comprise citizens and residents returning from across the globe. For example, 12 SARS-CoV-2 genomes from returnees to New Zealand from India who arrived on the same flight fell across at least four genomic lineages and comprised sequence divergence of up to 34 single nucleotide polymorphisms. This represented far more genomic mutations than was observed in New Zealand during the first outbreak in March – May 2020. Such a high level of diversity in just a small sample of positive cases from India suggest that the currently available genomic data fails to encompass the true diversity that existed locally let alone globally.

The re-emergence of COVID-19 into New Zealand in August 2020 exemplified one of the most complete genomic data sets for a specific outbreak compiled to date, comprising 81% of positive cases (145 of 179 total PCR positive cases). Real-time genomic sequencing quickly informed track and trace efforts to control the outbreak, setting New Zealand on track to eliminate the virus from the community for the second time. The rapid genome sequencing of positive samples provided confidence to public health teams regarding links to the outbreak and identified that cases and sub-clusters were linked to a single genomic lineage, resulting from a single introduction event. Indeed, the timing and length of lockdown measures were partly informed based on these data.

Nevertheless, the biased nature of global sampling, including the contribution of very few genome sequences from certain geographic locations, clearly limited the power of genomics to attribute the geographical origin of New Zealand’s August 2020 outbreak. We therefore advocate that careful consideration of the potential sampling biases and gaps in available genomic data be made whenever attempting to determine the geographic origins of a specific outbreak of SARS-CoV-2. Analysis should consider all available evidence, including from genomic and epidemiological sources.

## Methods

### Ethics statement

Nasopharyngeal samples testing positive for SARS-CoV-2 by real-time polymerase chain reaction (RT-PCR) were obtained from public health medical diagnostics laboratories located throughout New Zealand. All samples were de-identified before receipt by the researchers. Under contract for the New Zealand Ministry of Health, ESR has approval to conduct genomic sequencing for surveillance of notifiable diseases.

### Genomic sequencing of SARS-CoV-2: New Zealand’s second wave

A total of 172 (out of 179) laboratory-confirmed samples of SARS-CoV-2 were received by ESR for whole genome sequencing, comprising samples from New Zealand’s August 2020 outbreak. Genome sequencing of SARS-CoV-2 samples was performed as before^7^. In short, viral extracts were prepared from respiratory tract samples where SARS-CoV-2 was detected by RT-PCR using WHO recommended primers and probes targeting the E and N gene. Extracted RNA from SARS-CoV-2 positive samples were subject to whole genome sequencing following the ARTIC network protocol (V3) (https://www.protocols.io/view/ncov-2019-sequencing-protocol-v3-locost-bh42j8ye) and the Massey University 1200 bp primer set (https://www.protocols.io/view/ncov-2019-sequencing-protocol-rapid-barcoding-1200-bh7hj9j6)^21^.

Briefly, one of the tiling amplicon designs were used to amplify viral cDNA prepared with SuperScript IV. Sequence libraries were then constructed, using the Oxford Nanopore ligation sequencing and native barcoding expansion kit for samples amplified with the ARTIC V3 primer sets, and the Oxford Nanopore rapid barcoding kit for samples amplified with the 1200 bp primer sets. The 1200 bp primers and rapid barcoding was used in cases where the genomes were required urgently. Libraries were sequenced using R9.4.1 MinION flow cells, respectively. Near-complete (>90% recovered) viral genomes were subsequently assembled through reference mapping. Steps included in the pipeline are described in detail online (https://github.com/ESR-NZ/NZ_SARS-CoV-2_genomics). The reads generated with Nanopore sequencing using ARTIC primer sets (V3) were mapped and assembled using the ARTIC bioinformatics medaka pipeline (v 1.1.0). In total, 145 (out of 172) genomes from New Zealand’s August 2020 outbreak passed quality control. All data are available on GISAID.

### Phylogenetic analysis of SARS-CoV-2

All human SARS-CoV-2 genomes available on GISAID^4^ as of 25 September 2020 (n = 109,379) were downloaded and their lineages assigned according to the proposed nomenclature^16^ using pangolin (https://github.com/hCoV-2019/pangolin). Genomes assigned to the B.1.1.1. lineage (n = 4,544) were subsampled to include 1,996 most recent-in-time sequences along with four outgroup (non-B.1.1.1.) sequences, and were aligned with those from the recent New Zealand outbreak (n = 140) using MAFFT (v 7)^22^ employing the FFT-NS-2 progressive alignment algorithm. Bayesian phylogenetic analyses were performed using BEAST 2.5^23^. We used a strict clock model with one HKY substitution model (estimated frequencies) for each codon position and one for non-coding positions. We employed the Bayesian skyline model^24^ as a tree prior to allow effective population sizes to change over time intervals. These components of the model and their prior distributions are those used by Douglas et al. 2020^13^. Phylogenetic trees were annotated using FigTree (v 1.4)^25^ and Tree of Life (v 4)^26^.

## Data Availability

All data is available on GISAID database

https://www.gisaid.org/

## Acknowledgements

We thank the ARTIC network for making their protocols and tools openly available and specifically Josh Quick for sending the initial V1 and V3 amplification primers. We thank the diagnostic laboratories that performed the initial RT-PCRs and referred samples for sequencing as well as the public health units for providing epidemiological data. We thank Genomics Aotearoa for their support, the NextStrain team for their support and timely global and local analysis, and all those who have contributed SARS-CoV-2 sequences to GenBank and GISAID databases. The authors wish to acknowledge the use of New Zealand eScience Infrastructure (NeSI) high performance computing facilities. New Zealand’s national facilities are provided by NeSI and funded jointly by NeSI’s collaborator institutions and through the Ministry of Business, Innovation and Employment’s Research Infrastructure programme (https://www.nesi.org.nz).

