## Supplementary Figure 1 for "The power and limitations of genomics to track COVID-19 outbreaks: a case study from New Zealand"

### Supplementary Material

**Supplementary Figure 1**. Phylogenetic tree as presented in Figure 2a of the main article, although only taxa from the recent New Zealand outbreak are included, alongside five closest outgroup taxa. Clade posterior support values (percentages) are labelled on internal nodes. Alignment positions are only included in the figure if the site contains at least two variants among the taxa considered (ambiguous nucleotides are ignored from the count). The minority character in each column is coloured.

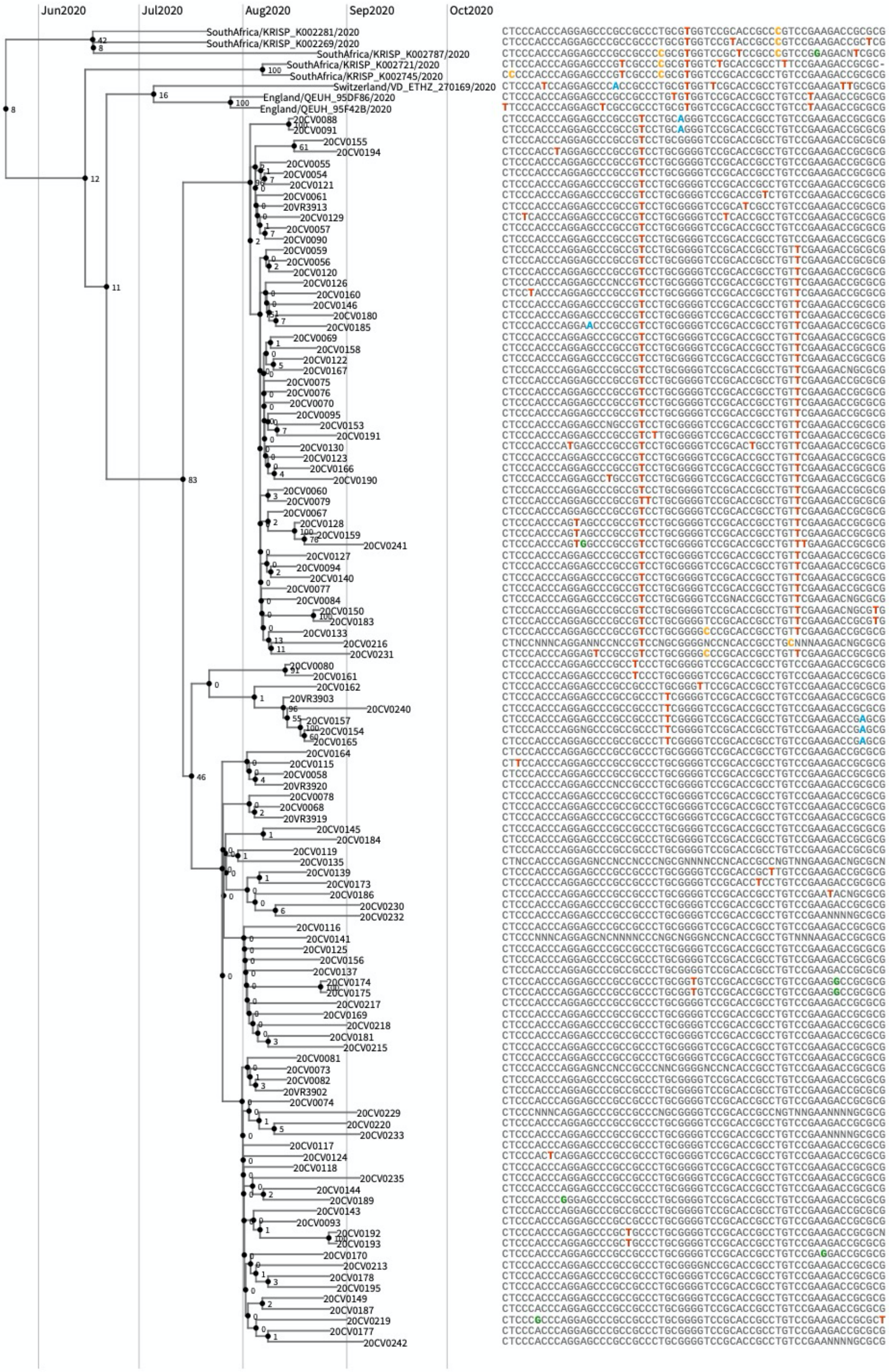
